# A Socio-spatial Model of the Risk of Hospitalization from Vulnerability to High Temperatures

**DOI:** 10.1101/2025.03.29.24319024

**Authors:** Juan Declet-Barreto, Benjamin L. Ruddell, Jarrett J. Barber, Diana B. Petitti, Sharon L. Harlan

## Abstract

Urban heat islands and climate change create increasingly hot environments that pose a threat to the health of the public in urban areas throughout the planet. In Maricopa County, Arizona, --- the hottest metropolitan area in the United States---we have previously shown that the effects of heat on mortality are greater in the social and built environments of low-income and communities of color (predominantly Hispanic/Latinx and Black neighborhoods).

In this analysis of morbidity data from Maricopa County, we examined the relationship between heat-related hospitalization and summertime daily maximum air temperatures in groups defined at the census block group level as being at high, medium, or low vulnerability based on a Heat Vulnerability Index that was derived from socio-economic and built-environment data.

For all three categories of census block group heat vulnerability, we identified 26°C as the daily maximum air temperature threshold beyond which heat-related hospitalization risk increased rapidly with each 1 °C increase in temperature. Compared to this baseline temperature, the relative risk of hospitalization was greatest in the high vulnerability census block groups and least in the low vulnerability census block groups with intermediate increases in the medium vulnerability census block groups. Specifically, with 26°C as the referent, the relative risks of heat-related hospitalization increased from 0.97 at 27°C to 15.71 at 46°C in the low vulnerability group, from 1.03 at 27°C to 53.97 at 46°C in the medium vulnerability group, and from 1.09 at 27°C to 162.46 at 46°C in the high vulnerability group.

Our research helps identify areas with high heat population sensitivity and exposure that can be targeted for adaptation with policies and investments, which include, for example, improving public health safety nets and outcomes, access to affordable energy-efficient housing and health care, energy justice, and modifications to cool the urban built environment. Our hospitalization risk estimates can be incorporated into quantitative risk assessments of heat-related morbidity in Maricopa County.

## Introduction

Strong relationships between high temperatures and human mortality (1–7) and morbidity (7–10) are well established around the world. In addition to heat-related illness and death directly caused by exposure, exreme heat elevates mortality and morbidity rates in people with cardiovascular and respiratory diseases, diabetes, and other chronic health conditions (6,11,12). A study that modeled climate change in 43 countries for the period 1991-2018 reported that anthropogenic climate change was responsible for 37% of warm season heat-related deaths (13). The Sixth Assessment Report of the Intergovernmental Panel on Climate Change (IPCC AR6) provides ample evidence that elevated temperatures pose ever greater risks to human health in the summer season as global temperatures rise and extreme heat events become more intense and frequent (14).

Against the backdrop of global temperature increases, cities are warming more than their surroundings due to the Urban Heat Island (UHI, 15). Urbanization-induced warming significantly contributes to regional warming independently of warming due to greenhouse gases (16–18). UHIs are formed by regional and local anthropogenic processes of rapid population growth, urban expansion, and land use/land cover changes (19). Changes in built environments exert pressures on natural ecosystems that are already under stress due to global climate change-induced alterations (20), and alter the spatial distribution of ecosystem services that regulate temperatures (21).

At the neighborhood scale, land covers -- the percentages of impervious surfaces and vegetated areas – are correlated with air temperatures, and consequently, heat stress in the population (4,22,23). Spatially heterogeneous microclimates in cities follow the pattern of land cover and land use (24,25). The magnitude of the UHI is typically greater where there are higher rates of solar energy absorption into vertical and horizontal impervious surfaces, denser concentrations of heat-emitting vehicular and industrial activities, and little cooling green space (26). Unequally distributed temperatures and disparities in ecosystem services that provide temperature reductions within cities are known to be highly correlated with neighborhood socio-economic characteristics (27–30) and heat-related mortality (31–33).

Household income is a predictor of sparse vegetation and dense impervious surfaces, two important drivers of intra-urban neighborhood differences in temperatures (34,35). Extensive impervious surfaces, lack of tree canopy and abundance of other heat-retaining land covers are more prevalent in predominantly African-American, Hispanic, and Asian neighborhoods than in whiter, more affluent areas (31,36,37). The spatial confluence of ecological and social hierarchies is underscored by research that shows unequal distributions of vegetated green spaces contribute to health disparities between low- and higher-income communities (38,39) and between population groups that are more or less sensitive to the harmful health effects of extreme temperatures (28).

In this study, we assess the risks of hospitalization attributed to extreme heat hazards in a physically and socio-economically diverse urban environment—Maricopa County, Arizona, USA, home to Phoenix and other sizeable municipalities – the hottest large metropolitan area in the United States. Specifically, we address the research question: what are the effects of high summertime temperatures on heat-related hospitalizations in neighborhoods with inequitable built environments and socio-economic conditions? Heat-related hospitalizations affect many times more people than heat-related deaths, are an indicator of human suffering, and are socially and financially costly. We estimate the relative risks of summertime heat-related hospitalizations as a function of maximum daily air temperature in geographically defined settings characterized by their vulnerability.

Our analysis is structured using concepts from the IPCC AR6 report (40), which provides a risk framework for assessing impacts of climate change hazards on individuals and communities resulting from interdependent human and ecological systems. *Vulnerability* is an important concept in health literature because it conveys the potential for environmental hazards to adversely affect populations. It is grounded in empirical evidence that unequal health and well-being outcomes occur along a socio-spatially differentiated spectrum of exposure, sensitivity, and adaptive capacity that can mitigate or exacerbate impacts (41–43). *Exposure* is defined here as “places and settings” (i.e., neighborhood built environments) that provide different levels of ecosystem services and infrastructure resources for residents (14,43). *Sensitivity,* studied extensively in the health field, denotes the degree to which different populations are susceptible to the deleterious effects of exposure to environmental hazards (14,43). These concepts are commonly used throughout the literature on human dimensions of climate change to describe vulnerability to extreme heat and a variety of other hazards (e.g., cold temperatures, flood, drought; see 44). To conclude our analysis, we suggest how our findings can be applied by policymakers who manage adaptations – the third component of vulnerability – to reduce adverse impacts of extreme heat on marginalized communities and promote social justice.

## Materials and Methods

### Study Area

Maricopa County, Arizona, is located in the Sonoran Desert of the southwestern United States. Phoenix, the county seat and state capital, has a summertime (June, July, August) mean daily maximum temperature of 40.4°C (45). Originally inhabited around the 14^th^ century by small communities of Indigenous people engaged in subsistence farming, the first white settlers arrived in Phoenix in the late 19^th^ century and developed large-scale, commercial agricultural enterprises (46,47). By the mid-twentieth century, Maricopa County had become one of the fastest-growing metropolitan regions in the United States, increasing from 90,000 persons in 1920 to 332,000 in 1950, to 3.0 million in 2000. In 2020, Maricopa County had a population of nearly 4.4 million people, who mostly live in the 25 self-governing municipalities centered around Phoenix (48).

Over the previous two decades before 2020, heat-associated deaths reported in Maricopa County increased progressively from 21 in 2001 to 645 in 2023, representing an accelerating trend in the years prior to 2020 (e.g., a large annual increase from 2019, when there were 199 heat-associated deaths in the county; see 49,50).

Urbanized central Arizona is an ideal place to study heat-related vulnerability given its explosive urban growth during the 20^th^ century, extremely high summertime temperatures, and large-scale land-use/land cover modifications of agricultural and desert landscapes (51,52). The city of Phoenix UHI is most intense in the central business district and temperatures generally diminish with increasing distance from there (53,54). At the regional scale, however, the decentralized pattern of urban development has led to the emergence of "mini-UHIs" in the central business districts of the larger municipalities that make up the Maricopa County urban area, creating much variability in UHI intensities throughout the metropolitan area (55).

### Heat-Related Hospitalizations

Records of heat-related inpatient hospital visits from 2005 through 2009 were extracted from databases of Maricopa County acute-care hospitals (excluding Veterans’ Administration, Indian Health Service, and military hospitals). The dataset used in this study was assembled by Arizona State University’s Center for Health Information and Research based on data submitted by hospitals to the Arizona Department of Health Services (ADHS). These data were available to the authors under IRB HS 11-0006 between ADHS and Arizona State University, and were accessed by authors approved by the IRB protocol on November 10, 2016 for research purposes. The authors did not have access to information that could identify individual participants during or after assembly of the dataset.

For the period 2005-2007, the data submitted to the ADHS included information on up to nine International Classification of Disease 9th Edition, Clinical Modification (ICD-9-CM) discharge diagnoses and up to four external causes of injury (E) codes. For the period 2008-2009, the data included information on ICD-9-CM discharge codes (<= 25) and external cause of injury (E) codes (<=9). Hospitalizations with the following ICD-9-CM codes as either a discharge diagnosis or as an external cause of injury in any position were used to identify conditions directly due to exposure to high environmental heat: 692.71, 692.72, 692.79, 708.8x, 992.xx, E904.2 (56).

For this analysis, we merged two inpatient datasets for 2005-2007 and 2008-2009, which were provided separately because the number of captured codes increased in 2008 from nine to 25 discharge codes and from four to nine E-codes. In an epidemiological or longitudinal research design, merging the two datasets would be incorrect because estimated changes in inpatient hospitalizations rates could reflect reporting changes in the number of captured codes and not changes in real rates of heat-related hospitalization. However, the change in number of captured codes is less of a concern in this study because all locations had the same number of possible codes in any given year, and we aggregated hospitalizations across all years in the five-year study period. Dates of hospital admission and discharge, patient demographic information, and latitude and longitude from the geocoded street address of a patient’s place of residence at the time of hospitalization were included in the dataset.

Hospitalizations considered in the analysis were further restricted as follows. First, to account for hospitalizations during periods of summertime temperatures, only those that occurred between May and October—corresponding to the Maricopa County surveillance period for heat-related deaths (57)—were selected. Only hospitalizations with a residential locator that identified the census block group (CBG) of residence or finer scale were selected. Of the 1,569 hospitalizations directly related to heat exposure, 160 were eliminated from the analysis because the date of admission was outside the summertime surveillance period, 218 were eliminated because the patient’s residence was outside of the county urban area, and 28 were eliminated because neither condition was met. From the remaining 1,163 eligible hospitalizations, 156 were excluded from the analysis because the residential locator identified only a zip code, census tract, or county. This left 1,007 hospitalizations (64.2 percent of all hospitalizations categorized as directly related to heat) for our analysis.

### Temperature

The heat hazard was characterized as daily maximum air temperature (*t_max_,*°C). Daily air temperature observations were obtained from 29 weather stations across three monitoring networks throughout Maricopa County (58–60). Only stations with continuous observations for May through October in each study year were used. Where necessary, observations were converted from Fahrenheit to Celsius and rounded to the nearest integer. Some stations outside the urban area were included in the study because they were the closest stations to many of the geocoded hospitalizations in the urban fringe. Table 1 summarizes monthly temperatures (May-October) recorded during 2005-2009 at the 29 county weather stations. Daily *t_max_* on the date of hospital admission was assigned from the nearest weather station to the CBG of residence of each of the 1,007 geocoded addresses.

**Table 1.** Monthly May-October Daily Minimum, Maximum, and Mean Air Temperatures Averaged for 29 Weather Stations Used in Maricopa County 2005-2009 Heat-Related Hospitalization Analysis.

| Month | Temperature (°C) |  |  |  |
| --- | --- | --- | --- | --- |
|  | Minimum | Maximum | Mean | SD |
| May | 12.8 | 46.1 | 34.4 | 4.6 |
| June | 25.0 | 48.9 | 39.0 | 3.8 |
| July | 26.1 | 50.0 | 40.8 | 3.3 |
| August | 26.7 | 47.2 | 39.3 | 3.1 |
| September | 23.3 | 47.8 | 36.8 | 3.2 |
| October | 11.1 | 41.1 | 30.5 | 4.2 |

### Vulnerability to Heat

Following previous methods (33,61), we prepared data to calculate a Heat Vulnerability Index (HVI). Using a Geographic Information System (GIS), we overlaid the 2010 Urban Areas boundary for Maricopa County (62) with CBG boundaries from the 2010 U.S. Census. We chose CBGs as spatial proxies for neighborhoods because they are subdivisions of census tracts and are typically socially and ecologically homogeneous (4,31). CBGs with centroids outside the Urban Areas boundary were excluded from the analysis. In addition, we eliminated four CBGs within the urban areas that we identified as containing institutional or transitory populations (i.e., college dormitories, state prison, and state hospital). Of the 2,505 CBGs in Maricopa County, the HVI was calculated for 2,387 (95.1 percent) in our analysis.

The measures of heat vulnerability (sensitivity and exposure) are the same as those used in previous research to construct an HVI to predict the socio-spatial distribution of heat-related mortality in Maricopa County for the same period (31), except that “not White” replaced two other measures of ethnicity (“Ethnic minority” and “Latino immigrant”). A justification for the selected population and land cover variables and a correlation matrix is provided in Supplemental Information, Section 1.

### Heat Vulnerability Index

The first step in calculating the HVI was a Principal Components Analysis (PCA) using all the measures of heat vulnerability shown in Table 2. Table 3 shows the results of the PCA. We applied varimax rotation to the PCA structure and three principal components were selected as factors. Variables correlated to socio-economic characteristics make up the first factor, labeled "Socioeconomic Sensitivity": percent Not White, percent No Air Conditioning (AC), percent No High School diploma, and percent Below Poverty. Percent Age 65 or Older, percent Living Alone, and percent Age 65 and Older Living Alone loaded into a second factor, labeled "Elderly/Living Alone Sensitivity". Finally, Unvegetated Surface mean and standard deviation loaded into a third factor labeled "Built Environment Exposure". The *Eigenvalues* for these three factors were all > 1.5. The total explained common variance was 79.0 percent.

**Table 2.** Measures of Health Outcome, Heat Hazard, Heat Vulnerability (Sensitivity and Exposure), and Population with Data Source, Time Period and Geographic Scale for Each Measure. Maricopa County 2005-2009 Heat-Related Hospitalization Analysis.

| Measure | Source | Time Period | Geographic Scale |
| --- | --- | --- | --- |
| **Health Outcome** |  |  |  |
| Heat-related Hospitalization | ADHS<sup>1</sup> | 2005-2009 | Point |
| **Heat Hazard** |  |  |  |
| Daily Maximum Air Temperature | AZMET, MFCDC, NOAA<sup>2</sup> | 2005-2009 | Point |
| **Heat Vulnerability** |  |  |  |
| <u>Sensitivity</u> |  |  |  |
| Not White | U.S. Census Bureau | 2010 | Census Block Group |
| No Air Conditioning | Maricopa County Assessor | 2010 | Parcel |
| No High School Diploma | U.S. Census Bureau | 2006-2010 | Census Block Group |
| Below Poverty | U.S. Census Bureau | 2006-2010 | Census Block Group |
| Age 65 or Older | U.S. Census Bureau | 2010 | Census Block Group |
| Living Alone | U.S. Census Bureau | 2010 | Census Block Group |
| Age 65 and Older Living Alone | U.S. Census Bureau | 2010 | Census Block Group |
| <u>Exposure</u> |  |  |  |
| Unvegetated Surface (Mean) | Landsat<sup>3</sup> | 2006 | 30 meters/pixel |
| Unvegetated. Surface (SD) | Landsat<sup>3</sup> | 2006 | 30 meters/pixel |
| **Population** |  |  |  |
| Number of Residents | U.S. Census Bureau | 2010 | Census Block Group |

1 Arizona Department of Health Services
2 Arizona Meteorological Network, Maricopa County Flood Control District, and National Oceanic and Atmospheric Administration
3 National Land Cover Dataset 2006 (Fry et al. 2011; reference 30)

**Table 3.** Results of Principal Components Analysis of Measures of Heat Vulnerability in 2,387 Maricopa County Census Block Groups. Maricopa County 2005-2009 Heat-Related Hospitalization Analysis.

| Measure | Factors and Factor Labels |  |  |
| --- | --- | --- | --- |
|  | Factor 1<br>Socioeconomic<br>Sensitivity | Factor 2<br>Elderly/<br>Living<br>Alone<br>Sensitivity | Factor 3<br>Built<br>Environment<br>Exposure |
| Not White | **0.85** | -0.36 | 0.09 |
| No Air Conditioning | **0.82** | 0.00 | 0.08 |
| No High School Diploma | **0.88** | -0.07 | 0.12 |
| Below Poverty | **0.83** | 0.02 | 0.12 |
| Age 65 or Older | -0.24 | **0.86** | -0.10 |
| Living Alone | 0.05 | **0.77** | 0.03 |
| Age 65 or Older and<br>Living Alone | -0.08 | **0.95** | -0.03 |
| Unvegetated Surface<br>(Mean) | 0.24 | 0.06 | **0.89** |
| Unvegetated Surface (SD) | 0.04 | -0.13 | **0.92** |

Bold values indicate measures that loaded together in the Principal Components Analysis

Next, a score for each of the three factors was calculated for each CBG in urban areas of Maricopa County by dividing each of the three factors into tertiles. Each CBG was then assigned a value of 1, 2, or 3 for each factor based on which tertile it fell into comparing the score to all of Maricopa County. For example, a value of 3 (more vulnerable) for Socioeconomic Sensitivity was given to a CBG when its score based on this factor’s socio-economic characteristics in that CBG was in the highest tertile. Similarly, a value of 1 (less vulnerable) for Socioeconomic Sensitivity was given to a CBG when its score in that CBG was in the lowest tertile relative to CBGs in urban areas of Maricopa County. The same was done for the other two factors, assigning values of 1, 2, or 3 for each factor to each CBG based on tertiles compared to all of Maricopa County, with values of 1 representing least vulnerable, and 3 the most vulnerable CBGs.

Next, for each CBG, the values for the three factors were summed to yield a summary HVI. The summary HVI could take values from 3 (least vulnerable considering Socioeconomic Sensitivity, Elderly/Living Alone Sensitivity, and Built Environment Exposure) to 9 (most vulnerable considering Socioeconomic Sensitivity, Elderly/Living Alone Sensitivity, and Built Environment Exposure). For the analysis, the CBGs were categorized based on the summary HVI by dividing into one of three categories: low HVI (summary HVI 3-5, n=410), medium HVI (summary HVI 6-7, n=1,573), or high HVI (summary HVI 8-9, n=400).

### Observed Rates of Heat-related Hospitalizations

A count of heat-related hospitalizations was observed in each CBG at daily maximum temperature (*t_max_*) along with the corresponding number of days for which that value of *t_max_* was observed in a CBG over the study period. Multiplying this number of days by a CBG’s population gives the number of person-days associated with the CBG’s heat-related hospitalizations observed at temperature *t*. Let *H_jt_* denote the sum of these heat-related hospitalization counts observed at temperature *t* over all CBGs in HVI group *j, j*=low, medium, high. Similarly, let *PD_jt_* denote the corresponding sum of person-days at temperature *t* over all CBGs in HVI group *j*.

Re-indexing from above, let *i(j,t)* map the *j*^th^ group at temperature *t* to the *i*^th^ unique combination of group and temperature. This results in n=64 observed combinations now denoted as *HVI_i_* and *t_i_*, i=1,…,n, where *HVI_i_* is a categorical variable taking one of the values of low, medium and high. Similarly, *H_i_* and *PD_i_* denote the corresponding counts and person-days, previously denoted as *H_jt_* and *PD_jt_*, respectively. Thus, the data used for our analysis consist of n=64 observations of *H_i_*, *PD_i_*, *t_i_* and *HVI_i_*. Corresponding observed heat-related hospitalization rates are *E_i_* = *H_i_*/*PD_i_*.

### Basic Model of Heat-related Hospitalization Rates

Our basic model of the mean, or expected, observed hospitalization rate, *H/PD*, as a function of *t* is

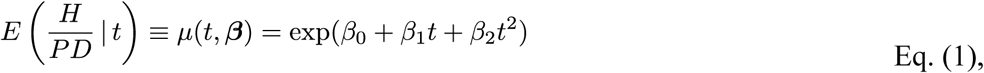

or, equivalently, the mean model of count, *H*, is

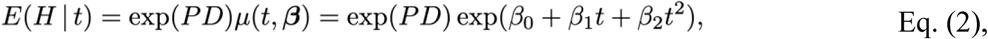

where *exp(PD)* is a multiplicative offset to adjust rates to a common per person-day scale. We adopted a quasi-Poisson model to account for over-dispersion relative to the Poisson distribution, thus retaining the maximum likelihood estimates of a Poisson rate model while better reflecting the variability of the data in our results. Customarily, we henceforth refer to rates as risks (see Supplemental Information Section 2 for detail about the models we evaluated).

We summarize the relative risks of heat-related hospitalization from a number of perspectives, including baseline relative risk for each HVI category using a baseline temperature of *t*_max_=26°C at which the overall relative risk begins to rise rapidly; relative risk of a one-degree increase in temperature for each HVI category; and relative risk of vulnerability to compare risk among HVI categories. Details sufficient to reproduce our risk model and our determination of the baseline temperature are provided in Supplemental Information Section 2.

## Results and Discussion

### Socio-spatial Variability in Vulnerability

Vulnerability to heat-related hospitalizations is unevenly distributed throughout Maricopa County. Figure 1 shows the spatial distribution of the HVI, classified as low, medium, or high vulnerability (in blue, yellow, and red, respectively). The high vulnerability areas in the central city contain Phoenix’s sparsely vegetated, low-income neighborhoods with many communities of color, and consequently, the highest socioeconomic sensitivity and built environment exposure. A thin band of high vulnerability is along the US-60 highway to the east. It passes through Tempe and Mesa, cities that contain similar but smaller pockets of high vulnerability near their downtown cores. Age-restricted retirement communities in the northwest edge of the metro area, along with other recently built areas north and east of Phoenix, have high proportions of vulnerable residents. Supplementary Information Section 1 presents a map for each individual factor of the HVI.

**Figure 1.**
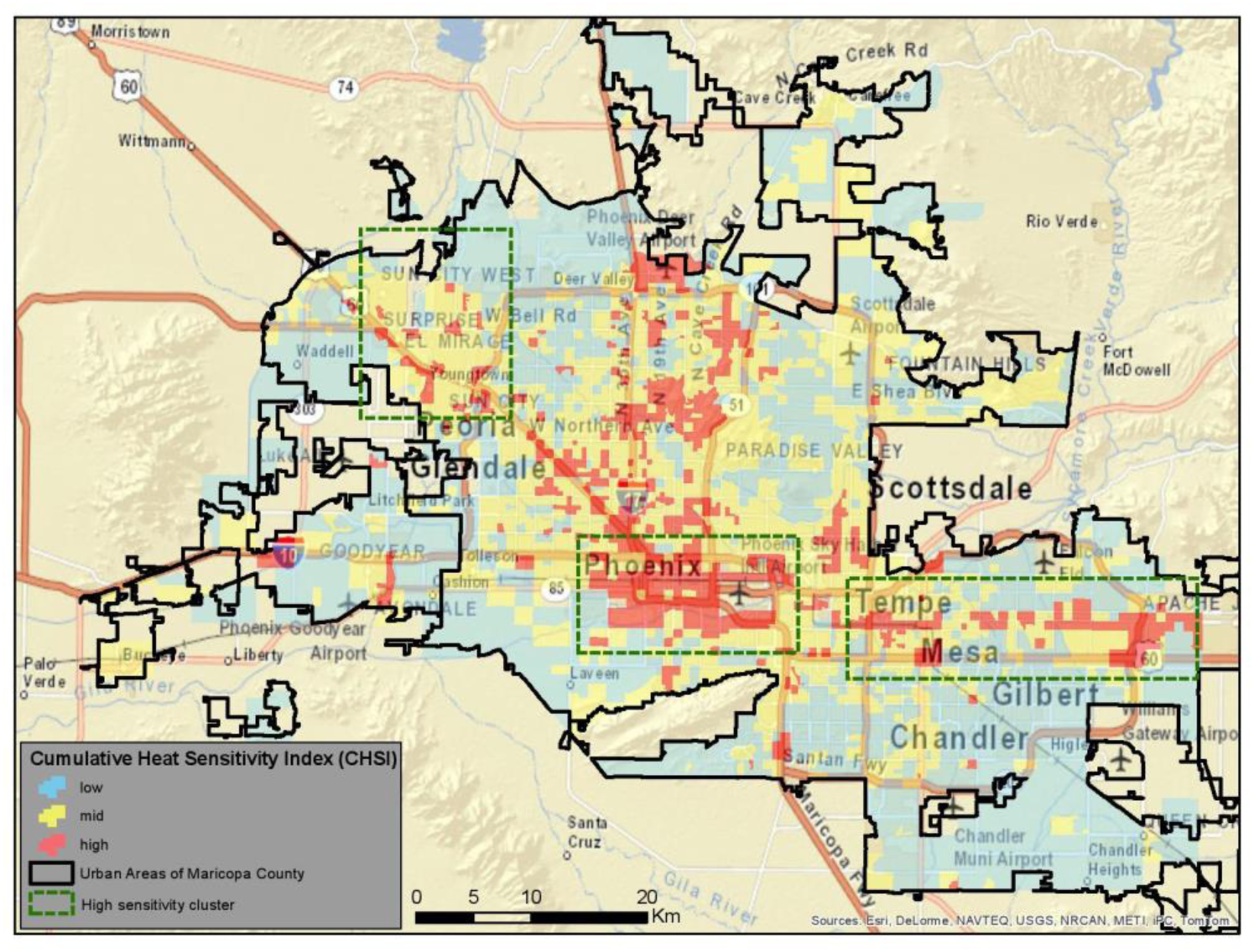
Heat Vulnerability Index in Census Block Groups in Maricopa County, AZ.

### Relative Risks of Heat-related Hospitalizations

For all HVI categories, the relative risk of hospitalization increases beyond a maximum daily air temperature *t*_max_=26°C; thus 26°C is our baseline temperature (Figure 2; see Supplementary Material, Section 2 for description of how the baseline temperature was determined). The relative risk of heat-related hospitalization increases rapidly with each 1°C increase in temperature (Table 4). Hospitalizations at *t*_max_ >46°C were observed, but relative risks were suppressed from Table 4 due to the small number of cases (see Supplementary Material Table S2 for a complete distribution of hospitalizations and person-days by *t*_max_). Overall, the relative risk increases from 1.03 at *t*_max_=27°C to 51.64 at *t*_max_=46°C; these estimates closely track the medium HVI category relative risk estimates.

**Figure 2.**
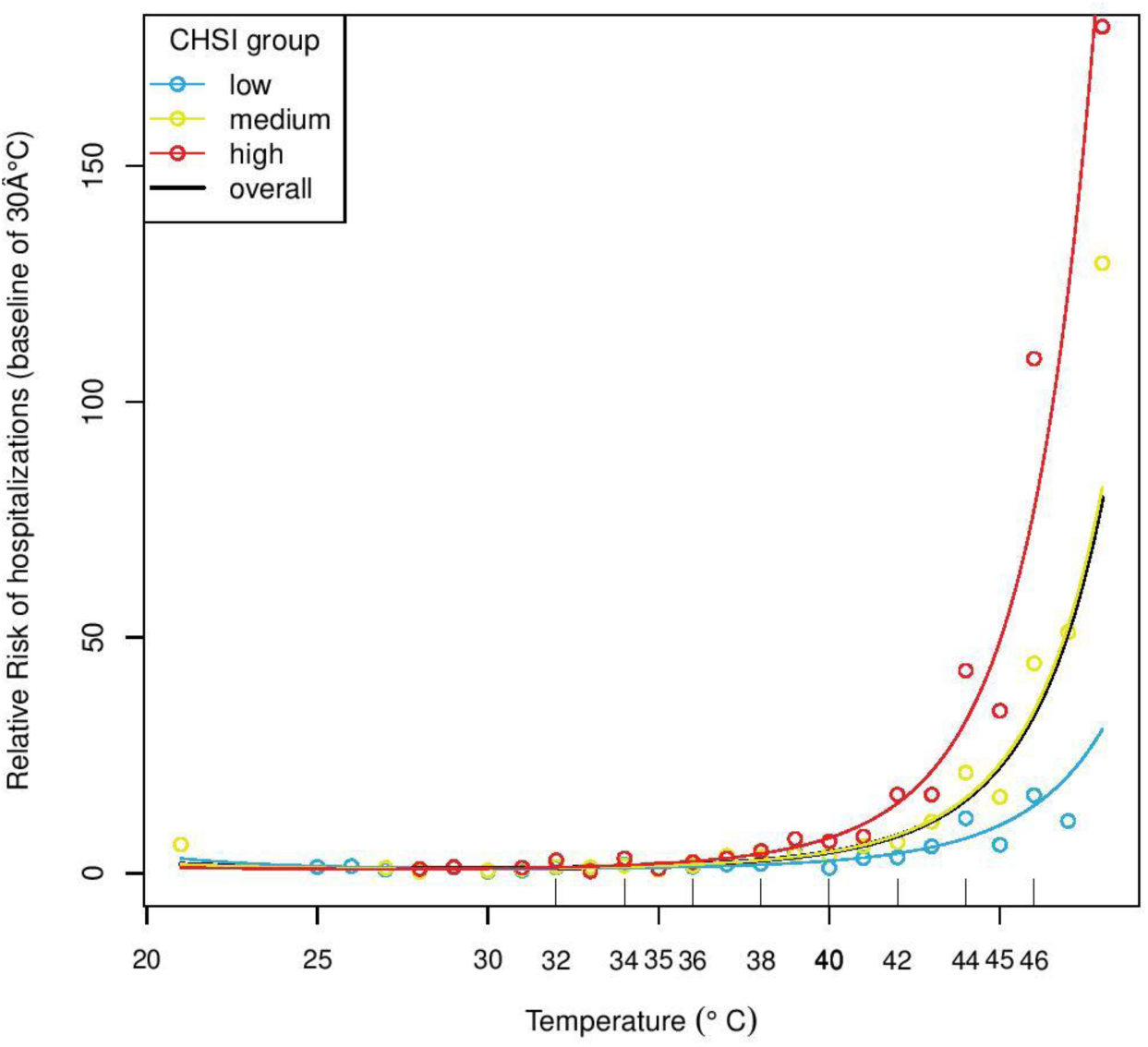
Relative risks of heat-related hospitalizations (baseline *t*_max_=26°C). HVI category risk curves are drawn using the same color scheme as in Figure 1.

**Table 4.** Relative Risk of Heat-Related Hospitalization and 95% Confidence Interval (CI) for Increasing Maximum Temperature (*t*_max_) by Category of Heat Vulnerability Index. Maricopa County 2005-2009 Heat-Related Hospitalization Analysis.

| $t_{\max}$<br>(°C) | Category of Heat Vulnerability Index | | | | | | | |
| --- | --- | --- | --- | --- | --- | --- | --- | --- |
|  | Low |  | Mid |  | High |  | Overall |  |
|  | Relative Risk | (95% CI) | Relative Risk | (95% CI) | Relative Risk | (95% CI) | Relative Risk | (95% CI) |
| 26 | referent | - | referent | - | referent | - | referent | - |
| 27 | 0.97 | (0.89-1.06) | 1.03 | (0.95-1.13) | 1.09 | (0.99-1.20) | 1.03 | (0.95-1.12) |
| 28 | 0.96 | (0.82-1.13) | 1.09 | (0.92-1.29) | 1.21 | (1.00-1.47) | 1.08 | (0.92-1.27) |
| 29 | 0.97 | (0.76-1.22) | 1.16 | (0.91-1.48) | 1.37 | (1.04-1.81) | 1.15 | (0.91-1.46) |
| 30 | 0.99 | (0.73-1.34) | 1.27 | (0.93-1.73) | 1.58 | (1.11-2.25) | 1.25 | (0.93-1.69) |
| 31 | 1.03 | (0.72-1.48) | 1.40 | (0.97-2.04) | 1.85 | (1.2-2.84) | 1.39 | (0.97-1.98) |
| 32 | 1.09 | (0.72-1.66) | 1.58 | (1.03-2.43) | 2.20 | (1.34-3.62) | 1.56 | (1.04-2.34) |
| 33 | 1.18 | (0.74-1.88) | 1.82 | (1.13-2.93) | 2.67 | (1.53-4.67) | 1.79 | (1.14-2.81) |
| 34 | 1.29 | (0.78-2.16) | 2.12 | (1.26-3.58) | 3.30 | (1.78-6.1) | 2.08 | (1.28-3.4) |
| 35 | 1.45 | (0.84-2.51) | 2.52 | (1.44-4.42) | 4.14 | (2.12-8.07) | 2.47 | (1.47-4.17) |
| 36 | 1.65 | (0.92-2.95) | 3.05 | (1.69-5.52) | 5.29 | (2.59-10.81) | 2.99 | (1.73-5.16) |
| 37 | 1.91 | (1.03-3.52) | 3.76 | (2.02-6.98) | 6.89 | (3.24-14.66) | 3.67 | (2.08-6.47) |
| 38 | 2.25 | (1.18-4.27) | 4.71 | (2.48-8.94) | 9.12 | (4.13-20.15) | 4.59 | (2.56-8.21) |
| 39 | 2.69 | (1.39-5.24) | 6.01 | (3.11-11.59) | 12.30 | (5.39-28.06) | 5.84 | (3.23-10.54) |
| 40 | 3.29 | (1.66-6.53) | 7.80 | (3.99-15.24) | 16.87 | (7.18-39.62) | 7.56 | (4.17-13.7) |
| 41 | 4.08 | (2.02-8.26) | 10.30 | (5.23-20.31) | 23.55 | (9.77-56.73) | 9.97 | (5.51-18.03) |
| 42 | 5.16 | (2.51-10.63) | 13.85 | (6.99-27.45) | 33.45 | (13.57-82.43) | 13.37 | (7.44-24.05) |
| 43 | 6.64 | (3.17-13.91) | 18.96 | (9.54-37.67) | 48.37 | (19.23-121.63) | 18.26 | (10.25-32.52) |
| 44 | 8.69 | (4.07-18.55) | 26.40 | (13.27-52.53) | 71.17 | (27.78-182.34) | 25.37 | (14.42-44.65) |
| 45 | 11.58 | (5.32-25.22) | 37.42 | (18.79-74.5) | 106.59 | (40.87-277.95) | 35.88 | (20.67-62.29) |
| 46 | 15.71 | (7.05-35) | 53.97 | (27.06-107.64) | 162.46 | (61.21-431.2) | 51.64 | (30.15-88.44) |

The relative risk of heat-related hospitalizations is higher for higher HVI categories when compared to lower HVI categories. In the low HVI category, the estimated relative risk of hospitalization increases from 0.97 at t_max_=27°C to 15.71 at t_max_=46°C, compared to that in the medium HVI category, increasing from 1.03 at t_max_=27°C to 53.97 at t_max_=46°C. The relative risk in the high HVI category is higher than in the low and medium HVI categories, increasing from 1.09 at t_max_=27°C to 162.46 at t_max_=46°C (Table 4; note that relative risk for t_max_=26°C is 1.00).

Our first finding is that the relative risk of heat-related hospitalizations in urban Maricopa County increases as daily maximum air temperature increases beyond a baseline temperature of 26°C. Second, the risk increases rapidly with each 1°C increase in temperature beyond the baseline. This finding is similar to a previous analysis of data on heat-related hospitalizations and emergency department visits in Maricopa County (9), where hospitalizations and emergency department visits were triggered at 27°C and 29°C daily maximum air temperature, respectively.

In our geographic analysis we characterized the urban population of Maricopa County according to vulnerability to the hazard of extreme heat. Our third finding is that the relative risk of heat-related hospitalization is higher in geographic areas that are more heat-vulnerable as measured using an index based on socioeconomic and age-related population variables and exposure based on the built environment. Further, higher population vulnerability groups have higher hospitalization risks than groups with lower population vulnerability even when controlling for temperature. For example, at the highest reported temperature for which reasonably reliable relative risks of hospitalizations could be estimated (46°C), the ratio of the relative risk of hospitalization in the high HVI category compared with the low HVI category was 10.3. Population heat risk factors mediate the effect of the heat hazard on health outcomes: less vulnerable population groups have lower hospitalization risks at a given temperature compared to those with higher vulnerability, as evidenced by the higher estimated relative risk of heat-related hospitalization in higher vulnerability groups.

Our fourth important finding is that three discrete factors correlate with risks of heat-related hospitalizations in neighborhoods: the spatial distribution of the HVI–constructed from socioeconomic, age and social isolation, and built environment factors–varies widely throughout urban areas, but in general, is highest in the Phoenix city urban core, lower in the expanding suburbs, and lowest near the suburban fringes. When the HVI is disaggregated into its factor components, however, a more nuanced pattern of heat risk emerges. The pattern is now one of high socio-economic sensitivity in the urban core among low-income communities of color with barren residential landscapes, and high sensitivity in certain areas of the suburban fringe with higher proportions of older people and people living in social isolation, also with low levels of vegetated land cover. Over the decades, residential and commercial development of the desert to the west and north of Phoenix has resulted in building some wealthy suburbs, but also considerable urban blight, spreading inequitable built environments across the urban fringe.

These two types of populations that are especially sensitive to adverse health effects of extreme heat –one made up of low-income communities of color in urban cores, the other suburban, white, and elderly–suggests a dual pattern of urban vulnerability that is not evident from looking only at cumulative vulnerability. Our findings are in line with previous research that has found higher rates of heat-related morbidity and mortality among lower income and elderly people in the United States (63–67) and in lower- and middle-income countries (7).

The HVI presented here validates the usefulness of a similar index used in an earlier study in Maricopa County based on year 2000 data (63). The two indices are similar in construction methodology and were evaluated using different health outcomes, and their findings are similar. Comparison of the indices against heat-related mortality and heat-related hospitalizations reveal that prediction of adverse health impacts coincides spatially with observed outcomes, suggesting that people of color living in densely built urban cores with low median income, and elderly individuals living in the desert suburban fringe, are at high risk for heat-related death or hospitalization. Both studies used variables similar to the index developed by Reid et al. (68) for U.S. counties (with the exception of diabetes-related morbidity among elderly persons), who later validated their index against heat-health outcomes, finding small but positive associations between their index and heat-related mortality and morbidity (61).

Another similar index for Phoenix developed by Chow et al. (2011) included only four socioeconomic (median income, old age, nativity, and residential mobility) and three biophysical (maximum and minimum temperature and NDVI, 69) variables. This index also identified urban core persons of color and suburban elderly individuals as at-risk for adverse heat-related outcomes. Chow et al. 2011 did not offer a nuanced explanation of socio-ecological characteristics that contribute to higher heat risks among heat-vulnerable groups, something that we attempted to do in our research.

There are several limitations of our study. The physiological and socio-demographic characteristics of individuals associated with elevated heat risks were scaled up to the census block group level to evaluate the relationship between exposure to high temperatures in geographically-defined areas, but they cannot entirely explain individual cases of heat-related hospitalizations. In a survey of individual households in four Phoenix study sites, Watkins et al. (70) found that CBG scores on Harlan et al.’s (63) HVI —which is very similar to the HVI presented here—were associated with differences in several household heat adaptive cooling behaviors, access to resources, and feeling of being too hot inside their home during the summer (a self-reported health outcome). However, for most of the difference effect sizes were small and there was considerable variation among households within CBGs. Research that combines data-rich, neighborhood-scale frameworks with individual assessments of heat exposure can help refine more equitable public health interventions to reduce the burden of heat on health in cities.

Another limitation of our study is that the administrative hospitalization data are gathered primarily to justify billing, and the assigned ICD codes may not exclusively reflect patients’ conditions (71). Furthermore, medical providers do not record conditions consistently and the mapping of ICD codes to conditions is sometimes not uniform. There is no standardization of the order of recording of conditions and ICD codes within an administrative record and reliance on the first code listed or only one code can lead to under-ascertainment of heat-related hospitalizations. We avoided the latter problem by selecting hospitalizations with a code related to heat in any position in a patient’s administrative record. This permitted us to capture co-morbidities exacerbated by extreme heat.

### Historical-geographical Production of Present-day Heat Vulnerabilities

We have identified contemporary markers of socio-ecological vulnerability to extreme heat through indicators such as poverty, race, age, isolation, and lack of vegetation, which may include trees and plants in yards, parks, and other public spaces. These are *manifestations* of vulnerability and not the *causal* factors that create vulnerability. To identify causal factors we must “shift the gaze to the historical and multi-causal production of harms” (72, 112) by framing present conditions within the historical trajectory of urban development in Maricopa County. Contemporary exposure to extreme heat in Maricopa County’s urban heat island has been created by the root causes of pro-growth urban development and racial discrimination (73). But the root causes of extreme heat vulnerability described here are not unique to Maricopa County, as they are found globally and in many cities across the United States, shaping population vulnerability in locally or regionally-specific ways. For example, the racist federal policy of residential redlining denied opportunities for wealth creation to people of color (but particularly African Americans) in hundreds of US cities, simultaneously increasing socio-economic sensitivity, decreasing access to coping resources among such groups, and increasing temperatures in formerly redlined areas (74,75). In Phoenix, although historically redlined neighborhoods cover a relatively small area around downtown (76), these areas have some of the highest land surface temperatures in the city (77).

The previously redlined, present-day environmental justice communities bear large burdens of exposure not only to climate-augmented extreme weather threats, but also to technological and industrial hazards (78–80). This social and spatial pattern is typical of older cities with traditional central business districts and post-industrial legacies (81,82), but it is also found in the Phoenix core (83). Our analysis reveals a second pattern of socio-spatial disadvantage that has not received as much attention. Through the sustained, large-scale mid-20^th^ century expansion of Maricopa County’s urban footprint, and the increased magnitudes and duration of heat waves exacerbated by local UHIs and global climate change, age- and social isolation-based vulnerabilities are now evident at the suburban fringes and also in second-tier urban cores. For example, in areas such as the urbanizing area north of the Phoenix core, Home-Owner Associations (HOAs) have placed restrictions on residential water use and favored xeriscape vegetation (84), potentially increasing built environment exposure to extreme heat. Large commercial enterprises, low-quality (but affordable) housing, drug and alcohol treatment centers and shelters, and assisted living facilities built in the desert fringe have created some unattractive environments that draw people with low income and education who need affordable homes and social services.

Extreme heat risks cut across many different determinants of human health and socio-economic wellbeing. Thus risk-reduction solutions to adapt to higher temperatures that endanger population health need to include, for example, improving public health safety nets and outcomes, access to affordable energy-efficient housing and health care, energy justice, and modifications to cool the urban built environment. These transformations should happen within participatory governance frameworks in which broad sectors of civil society, including impacted communities, scientists, and policy makers collaborate in the search for equitable solutions. In this regard, governments at all levels should align heat-protective resources, measures, and policies towards neighborhoods with high heat sensitivity, targeting vulnerable areas for public and private infrastructure investments, information campaigns, and social services.

Our insights are useful to align state and local government adaptation pathways that reduce the public health burden of extreme heat disease, but that also address social and historical inequities that have contributed to developing extreme heat vulnerabilities over time. For example, vegetation (or “cool”) corridors have recently been proposed in urban areas to create shaded and walkable paths to protect people from heat in their daily activities to and from the spaces of work, schooling, and leisure (85). Heat-reducing cool roofs (86), pavement (87) that reflects solar radiation, and green roofs covered in vegetation (88) are especially effective for lowering temperatures in densely built low-income areas. To identify target areas for public health interventions, our estimates of the relative risk of hospitalizations with respect to a 1°C increase in daily maximum air temperature can be incorporated as an exposure-morbidity response to estimate heat-related hospitalizations using the health impact function of the Environmental Protection Agency (EPA) Benefits Mapping and Analysis Program (BenMAP) model (89).

## Conclusions

Our study developed a socio-spatial model of hospitalizations from exposure and sensitivity to high temperatures that links current disparities in heat-related health outcomes to the social and ecological conditions of neighborhoods and the populations that reside in them. The interdisciplinary research framework we deployed here can help devise solutions to socio-spatial inequalities that shape the vulnerability of populations to climate-augmented extreme weather events in urbanized landscapes. As cities develop climate action plans and heat mitigation policies with strong social equity components, assessing fine-scale intra-urban differences in both heat exposure and population sensitivity will be critical in order to devise socially equitable policies that can reduce inequities in the burden of heat-related illness. Our research can help identify areas with high population sensitivity and heat exposure that should be targeted by policies and investments.

## Data Availability

The terms of the IRB agreement do not permit the authors to share these personally identifiable data.

https://www.dropbox.com/scl/fi/s03apfvschzqpqeo1bz9q/Supplemental-Information-Section-1-2-and-3-MedRxiv-FNL.pdf?rlkey=xs00r04pjrf5h7rpbz01c8khp&st=7x2w3cub&dl=0

## Ethics Statement

The authors declare that all relevant ethical guidelines have been followed, all necessary IRB and ethics committee approvals have been obtained, all necessary patient consent has been obtained and the appropriate institutional forms archived.

## Data Availability

Data on heat-related inpatient hospital case counts used in this research were obtained from Arizona State University’s Center for Health Information and Research based on data submitted by hospitals to the Arizona Department of Health Services (ADHS). These data were available to the authors based on an IRB-approved agreement (HS 11-0006) between ADHS and Arizona State University. The terms of the IRB agreement do not permit the authors to share these personally identifiable data.

