## Supplemental Material for "A Socio-spatial Model of the Risk of Hospitalization from Vulnerability to High Temperatures"

#### Supplemental Information. Section 1. Details on Derivation of the Heat Vulnerability Index (HVI)

We constructed the Heat Vulnerability Index (HVI) as an indicator of population sensitivity and exposure to heat-related hospitalizations and partitioned the population into one of three HVI groups of low, medium, and high vulnerability. T o answer our research question, we analyzed the effects of HVI group on heat-related hospitalization risks at different temperature thresholds.

Previous research labels this type of indicator a "heat vulnerability index" **(e.g., 1,2,3, others)**. In North American cities, research on the health impacts of extreme heat consistently finds that people living with low incomes, and people with limited access to air conditioning (AC) – many of whom are people of color – are more sensitive to heat and they experience worse adverse heat-related health outcomes **(4,5)**. Both the physical presence of an air conditioning (AC) unit in a dwelling and the ability to pay for its use are related to socio-economic status **(6,7)**. AC can provide immediate respite from heat exposure and can significantly reduce heat illness and death. Physiologically, the elderly are more sensitive to temperature and have elevated risk for heat-related mortality and morbidity **(8,9)**. Many elderly people live alone, resulting in social isolation, which contributes to poor physical and mental health outcomes generally. Social isolation is related to heat-related mortality **(10–12)**.

These sensitivity variables have been found together and in various combinations in Houston **(13)**, New York **(14)**, Philadelphia **(15)**, Toronto **(16,17)**, Chicago **(18,19)**, Detroit **(20)**, Phoenix **(3,21)**, and in areas across Massachusetts **(22)**. In other countries, research in Birmingham, United Kingdom **(23)** found highest risks for populations in city centers, while in Australian cities, Loughnan et al. **Error! Reference source not found.** reported that suburban locations where the elderly typically live, as well as central city neighborhoods made up of primary speakers of languages other than English are overburdened with heat risks. In Germany, Lissner et al. **(25)** found that the elderly living in areas of highest UHI intensity are most vulnerable. Tran et al. **(26)** found that heat exposure among urban slum dwellers in Ahmedabad, India was driven by occupation; and in Maricopa County, Arizona, heat-caused and heat-related mortality was higher among persons in outdoors occupations and those experiencing homelessness **(27)**.

Variables from the 2010 U.S. Census and American Community Survey Five-Year Estimates for 2006-2010 **(28,29)** were used to characterize the socio-spatial sensitivity of the residential population. Individual parcel data from the county cadastral registry **(30)** were used to calculate an AC deprivation indicator for CBGs that represents the percentage of single-family homes without central AC or evaporation cooling (swamp cooler).

Our measure of exposure in the built environment is the Unvegetated Surface indicator, derived by first calculating the Normalized Difference Vegetation Index **(NDVI, 31)** for neighborhoods based on a Landsat scene **(32)**. NDVI range is ±1.0, with positive values indicating more vegetation. NDVI mean and standard deviation were calculated, the means were multiplied by minus one to rescale the indicator in increasing order of sensitivity, and we label this indicator Unvegetated Surface. The table below shows that the variables selected for HVI are significantly correlated with each other.

**Supplemental Table S1. Mean, Standard Deviation (SD), and Pearson Correlation Coefficient for Measures of Heat Vulnerability in Census Block Groups (n = 2,387). Maricopa County 2005-2009 Heat-Related Hospitalization Analysis.**

| | Not White | No Air Conditioning | No High School Diploma | Below Poverty | Age 65 or Older | Living Alone | Age 65 or Older and Alone | Unvegetated Surface (Mean) | Unvegetated Surface (SD) |
|---|---|---|---|---|---|---|---|---|---|
| Mean | 39.9 | 10.3 | 15.3 | 14.0 | 14.1 | 25.2 | 8.0 | 21.9 | 9.4 |
| SD | 26.4 | 19.0 | 16.3 | 14.7 | 17.3 | 13.8 | 9.2 | 9.2 | 4.0 |
| Not White | | | | | | | | | |
| No Air Conditioning | 0.62*** | | | | | | | | |
| No High School. Diploma | 0.78*** | 0.65*** | | | | | | | |
| Below Poverty | 0.64*** | 0.57*** | 0.63*** | | | | | | |
| Age 65 or Older | -0.51*** | -0.19*** | -0.21*** | -0.22*** | | | | | |
| Living Alone | -0.21*** | 0.02 | -0.12*** | 0.10*** | 0.40*** | | | | |
| Age 65 or Older and Alone | -0.38*** | 0.08*** | -0.10 | -0.09*** | 0.87*** | 0.62*** | | | |
| Unvegetated. Surface (Mean) | 0.29*** | 0.22*** | 0.31*** | 0.29*** | -0.08*** | 0.06*** | -0.01 | | |
| Unvegetated. Surface (SD) | 0.14*** | 0.16*** | 0.17*** | 0.14*** | -0.21*** | -0.07*** | -0.14*** | 0.69*** | |

*p ≤ 0.05. **p ≤ 0.01. ***p ≤ 0.001.

These variables were entered into a Principal Components Analysis and three factors were derived. Table 3 in the main text shows the results of our Principal Components Analysis and Figure 1 in the main text maps the distribution of the total HVI scores for all three factors. The figures below map each of the three factors in the HVI: a) Socioeconomic Sensitivity, (b) Elderly/Living Alone Sensitivity, and (c) Built Environment Exposure.

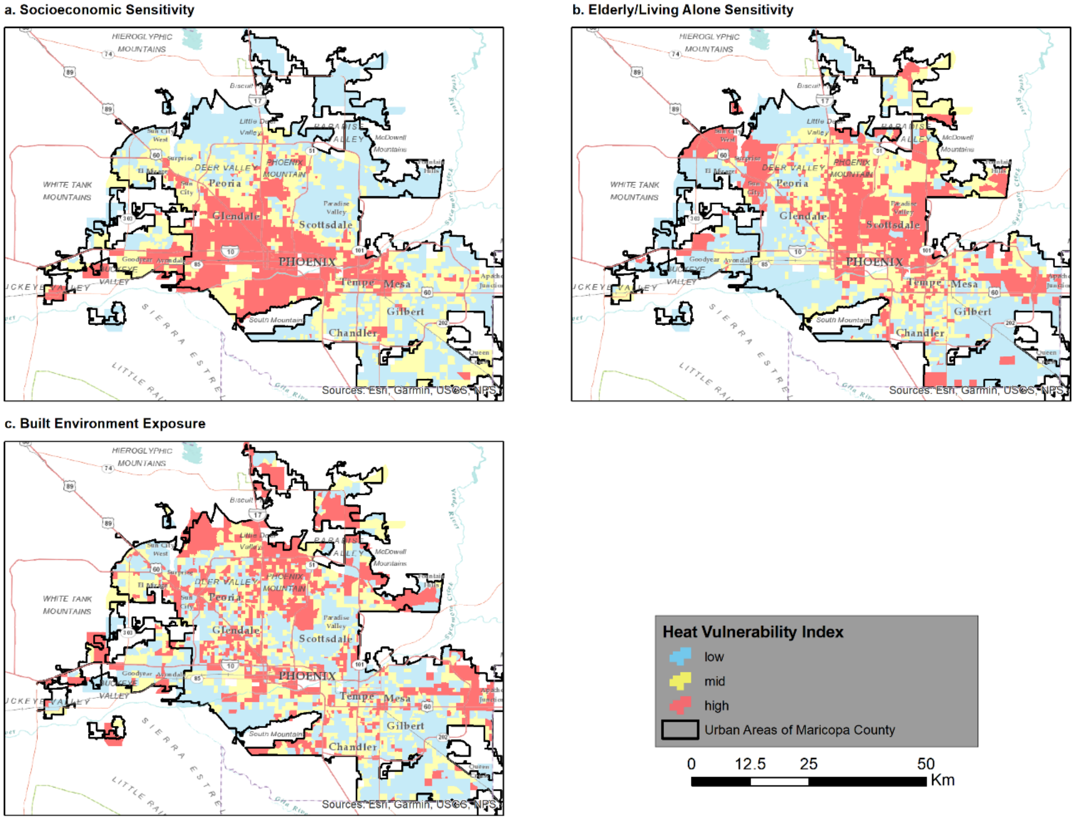


Supplemental Figure S1. Component Factors of Heat Vulnerability Index in Census Block Groups in Maricopa County, AZ.

A Socio-spatial Model of the Risk of Hospitalization from Vulnerability to High Temperatures

### Supplemental Information. Section 2. Details on Model Fitting

#### Overview

This section provides details on our model fitting of the heat-related hospitalizations data that served as rationale for choosing a quasi-Poisson distribution. Model fitting was conducted in R and code is reproduced below.

#### Model Fitting

The hospitalization rate data exhibit extra-Poisson dispersion, so we use maximum quasi-likelihood as implemented in R. We consider a sequence of four nested models of increasing complexity. The first model (Model 1) is our basic model (Eq. 1 in the main text), common to all HVI groups. The second model (Model 2) extends Model 1 to allow the HVI groups to have separate intercept parameters on the log scale for low, medium, and high HVI categories, respectively,

$$\mu_2(t, l=1, m=0, \boldsymbol{\beta}) = \exp((\beta_0+\beta_3)+\beta_1 t+\beta_2 t^2)$$
$$\mu_2(t, l=0, m=1, \boldsymbol{\beta}) = \exp((\beta_0+\beta_4)+\beta_1 t+\beta_2 t^2)$$
$$\mu_2(t, l=-1, m=-1, \boldsymbol{\beta}) = \exp((\beta_0+\beta_9)+\beta_1 t+\beta_2 t^2),$$

Note that Model 2 allows curves across HVI groups to differ only by a multiplicative constant, e.g., the medium HVI curve is obtained from the low HVI curve as

$$\mu_2(t, l=0, m=1, \boldsymbol{\beta}) = \exp(\beta_4-\beta_3)\mu_2(t, l=1, m=0, \boldsymbol{\beta}) .$$

Model 3 extends Model 2 to allow each HVI group to also have their own parameters associated with t,

$$\mu_3(t, l=1, m=0, \boldsymbol{\beta}) = \exp((\beta_0+\beta_3)+(\beta_1+\beta_5)t+\beta_2 t^2)$$
$$\mu_3(t, l=0, m=1, \boldsymbol{\beta}) = \exp((\beta_0+\beta_4)+(\beta_1+\beta_6)t+\beta_2 t^2)$$
$$\mu_3(t, l=-1, m=-1, \boldsymbol{\beta}) = \exp((\beta_0+\beta_9)+(\beta_1+\beta_{10})t+\beta_2 t^2),$$

Model 4 extends Model 3 to allow each group to also have their own parameters associated with $t^2$. Quasi-likelihood ratio tests indicate Model 2 performs significantly better than Model 1 and that Model 3 performs significantly better than Model 2, but that Model 4 does not perform significantly better than Model 3. Thus we choose Model 3 for our analysis in the main text. See R code and output that follows.

```
###############
rm(list=ls())
library(mgcv)

## Loading required package: nlme

## This is mgcv 1.8-33. For overview type 'help("mgcv-package")'.
```

```r
heat4.df<- read.csv(file="heat_hospitalizations_data.csv")
colnames(heat4.df)[1] = "tmax"
H<- c(heat4.df[,"H.low"], heat4.df[,"H.mid"], heat4.df[,"H.high"])
##length(H)
PD<- c(heat4.df[,"PD.low"],heat4.df[,"PD.mid"],heat4.df[,"PD.high"])
##length(PD)
Tmax<- rep(heat4.df[,"tmax"], 3)
##length(Tmax)

## Make HVI a factor (categorical variable):
HVI<- factor(rep(c("L","M","H"), c(28,28,28)), levels=c("L","M","H"))
## Next implements (log scale) effects with sum-to-zero contraints to
## facilitate ``overall'' risk, overall baseline risk and overall
## relative risk interpretation.
contrasts(HVI)<- contr.sum(levels(HVI))

heat.df<- data.frame(H=H,PD=PD,Tmax=Tmax,Tmax26=Tmax-26,HVI=HVI)

## Model 1 omits all HVI group specific parameters and fits a single,
##basic curve for all groups
quasiPoisHVIfitM1<- gam(H ~ offset(log(PD)) + Tmax26 + I(Tmax26^2),
                        family=quasipoisson(link=log), data=heat.df)
summary(quasiPoisHVIfitM1)

##
## Family: quasipoisson
## Link function: log
##
## Formula:
## H ~ offset(log(PD)) + Tmax26 + I(Tmax26^2)
##
## Parametric coefficients:
##                 Estimate Std. Error t value Pr(>|t|)
## (Intercept) -16.153076   0.458882 -35.201  < 2e-16 ***
## Tmax26        0.016110   0.072675   0.222  0.82513
## I(Tmax26^2)   0.008853   0.002888   3.066  0.00295 **
## ---
## Signif. codes:  0 '***' 0.001 '**' 0.01 '*' 0.05 '.' 0.1 ' ' 1
##
##
## R-sq.(adj) =  0.581   Deviance explained = 69.2%
## GCV = 4.6916  Scale est. = 4.3243    n = 84

## Model 2, allows different intecepts (log scale)
quasiPoisHVIfitM2<- gam(H ~ offset(log(PD)) + Tmax26 + I(Tmax26^2) + HVI,
                        family=quasipoisson(link=log),
                        data=heat.df)
summary(quasiPoisHVIfitM2)

##
## Family: quasipoisson
## Link function: log
##
## Formula:
## H ~ offset(log(PD)) + Tmax26 + I(Tmax26^2) + HVI
```

```
##
## Parametric coefficients:
##               Estimate Std. Error t value Pr(>|t|)
## (Intercept) -16.092238   0.292651 -54.988  < 2e-16 ***
## Tmax26        0.006390   0.046270   0.138    0.891
## I(Tmax26^2)   0.009440   0.001842   5.126 2.05e-06 ***
## HVI1         -0.677222   0.068046  -9.952 1.36e-15 ***
## HVI2          0.050817   0.056555   0.899    0.372
## ---
## Signif. codes:  0 '***' 0.001 '**' 0.01 '*' 0.05 '.' 0.1 ' ' 1
##
##
## R-sq.(adj) =  0.888   Deviance explained = 88.2%
## GCV = 1.8987  Scale est. = 1.7894    n = 84
```

```
## Model 3, additionally, allows different t terms
quasiPoisHVIfitM3<- gam(H ~ offset(log(PD)) + Tmax26 + I(Tmax26^2) + HVI +
                          HVI:Tmax26,
                        family=quasipoisson(link=log),
                        data=heat.df)
summary(quasiPoisHVIfitM3)
```

```
##
## Family: quasipoisson
## Link function: log
##
## Formula:
## H ~ offset(log(PD)) + Tmax26 + I(Tmax26^2) + HVI + HVI:Tmax26
##
## Parametric coefficients:
##               Estimate Std. Error t value Pr(>|t|)
## (Intercept) -16.173263   0.286151 -56.520  < 2e-16 ***
## Tmax26        0.021512   0.044712   0.481 0.631801
## I(Tmax26^2)   0.008785   0.001772   4.958 4.13e-06 ***
## HVI1          0.174407   0.240086   0.726 0.469774
## HVI2          0.027227   0.214595   0.127 0.899368
## Tmax26:HVI1  -0.059510   0.016456  -3.616 0.000531 ***
## Tmax26:HVI2   0.002206   0.014413   0.153 0.878739
## ---
## Signif. codes:  0 '***' 0.001 '**' 0.01 '*' 0.05 '.' 0.1 ' ' 1
##
##
## R-sq.(adj) =  0.894   Deviance explained = 90.2%
## GCV = 1.6589  Scale est. = 1.5876    n = 84
```

```
## Model 4, additionally, allows different t^2 terms
quasiPoisHVIfitM4<- gam(H ~ offset(log(PD)) + Tmax26 + I(Tmax26^2) + HVI +
                          HVI:Tmax26 + HVI:I(Tmax26^2),
                        family=quasipoisson(link=log),
                        data=heat.df)
summary(quasiPoisHVIfitM4)
```

```
##
## Family: quasipoisson
## Link function: log
```

```
##
## Formula:
## H ~ offset(log(PD)) + Tmax26 + I(Tmax26^2) + HVI + HVI:Tmax26 +
##     HVI:I(Tmax26^2)
##
## Parametric coefficients:
##                      Estimate Std. Error t value Pr(>|t|)
## (Intercept)        -1.612e+01  2.983e-01 -54.053  < 2e-16 ***
## Tmax26              1.558e-02  4.687e-02   0.332    0.741
## I(Tmax26^2)         8.948e-03  1.858e-03   4.816 7.44e-06 ***
## HVI1               -3.598e-02  4.028e-01  -0.089    0.929
## HVI2               -2.133e-02  3.823e-01  -0.056    0.956
## Tmax26:HVI1        -1.912e-02  6.561e-02  -0.291    0.772
## Tmax26:HVI2         7.862e-03  6.009e-02   0.131    0.896
## I(Tmax26^2):HVI1 -1.684e-03  2.688e-03  -0.626    0.533
## I(Tmax26^2):HVI2 -1.513e-04  2.379e-03  -0.064    0.949
## ---
## Signif. codes:  0 '***' 0.001 '**' 0.01 '*' 0.05 '.' 0.1 ' ' 1
##
##
## R-sq.(adj) =  0.892   Deviance explained = 90.2%
## GCV = 1.7366  Scale est. = 1.611     n = 84

beta<- dummy.coef(quasiPoisHVIfitM3)
anova(quasiPoisHVIfitM1,quasiPoisHVIfitM2, quasiPoisHVIfitM3,quasiPoisHVIfitM4
, test="LRT")

## Analysis of Deviance Table
##
## Model 1: H ~ offset(log(PD)) + Tmax26 + I(Tmax26^2)
## Model 2: H ~ offset(log(PD)) + Tmax26 + I(Tmax26^2) + HVI
## Model 3: H ~ offset(log(PD)) + Tmax26 + I(Tmax26^2) + HVI + HVI:Tmax26
## Model 4: H ~ offset(log(PD)) + Tmax26 + I(Tmax26^2) + HVI + HVI:Tmax26 +
##     HVI:I(Tmax26^2)
##   Resid. Df Resid. Dev Df Deviance  Pr(>Chi)
## 1        81     366.45
## 2        79     141.07  2  225.377 < 2.2e-16 ***
## 3        77     117.09  2   23.976 0.0005865 ***
## 4        75     116.29  2    0.805 0.7789181
## ---
## Signif. codes:  0 '***' 0.001 '**' 0.01 '*' 0.05 '.' 0.1 ' ' 1

## Parameter estimates
sum(beta$HVI) ## our sum-to-zero parameterization check

## [1] 0

sum(beta$"HVI:Tmax26") ## our sum-to-zero parameterization check

## [1] 0
```

##### Baseline Temperature

An intuitive feature of the risk curve $\mu(t, \beta)$ is the temperature $t$ beyond which risk begins to rise rapidly as temperatures increase. We refer to this as our baseline temperature. For our data, $\mu(t, \beta)$ is relatively constant up to this temperature, increasing rapidly thereafter, and below which the risk of hospitalization is presumably dominated by other risk factors beyond the scope of this study. While we do not know *a priori* what the value of this relatively constant baseline risk is, it is reasonable to expect that the first derivative of the risk curve, $\mu'(t, \beta)$, should be zero when approaching the region of rapidly increasing risk with increasing temperatures. T hus, for each HVI group, we use the temperature at which the first derivative of the risk curve increases above zero with increasing temperatures to indicate the baseline temperature. We now detail the process to estimate $\mu'(t, \beta)$ and to determine baseline temperature.

While the quasi-Poisson Generalized Linear Model (GLM) used throughout the main text provides a good fit to our data, we used a generalized additive model (GAM) with a reduced-rank thin-plate smooth spline linear predictor for estimating derivatives (33) as implemented in the R package, mgcv, shown in subsequent code and output. As we show below, the fit of a quasi-Poisson generalized additive model (GAM) is essentially the same as the quasi-Poisson GLM (Model 3), but we are more confident in the ability of such flexible GAMs to capture the local behavior of risk near the rapid rise in risk observed with increasing temperatures, and hence to identify a baseline temperature; the more familiar parametric linear predictors of our quasi-Poisson GLM (Model 3) and similar models are easier to interpret as we have discussed above and in the main text.

##### Quasi-Poisson GAM

A rule-of-thumb, when inferring about a function, using a smooth model, is that the model be continuous, also with continuous first and second derivatives. Therefore, to infer about the first derivative function of risk, we fit a risk function whose third derivative is continuous. As we should expect, the summary statistics of the smooth (reduced-rank thin plate) spline GAM fit here is very similar to that of our quasi-Poisson GLM (Model 3) used throughout the main text, but, again, we are more confident in the quasi-Poisson GAM to better capture the behavior of the derivative of the risk curve near the baseline temperature, and we prefer the interpretation of our quasi-Poisson GLM, as we have discussed above and in the main text. The code and output, below, fits the GAM and compares it to the GLM (Model 3); we see that the difference in deviance explained is small compared to the difference in degrees of freedom.

```
options(contrasts= rep("contr.trmt",2))
contrasts(heat.df$HVI)<- contr.treatment(levels(heat.df$HVI))
## Reduced rank thin plate regression splines used by default for each
## group after giving each group its own ``intercept.''  M=3 ensures
## use of model with continuous 3rd derivative.
quasiPoisgamHVIfit<- gam(H ~ offset(log(PD)) + HVI + s(Tmax26, m=3, by=
HVI),
                         family=quasipoisson(link=log),
                         data=heat.df)
## Fit of GAM is very similar to GLM used throughout main text:
summary(quasiPoisgamHVIfit) ## GAM

##
## Family: quasipoisson
## Link function: log
##
```

```
## Formula:
## H ~ offset(log(PD)) + HVI + s(Tmax26, m = 3, by = HVI)
##
## Parametric coefficients:
##             Estimate Std. Error t value Pr(>|t|)
## (Intercept) -15.1905     0.2168 -70.056   <2e-16 ***
## HVIM          0.4023     0.3137   1.283   0.2037
## HVIH          0.7381     0.4050   1.823   0.0724 .
## ---
## Signif. codes:  0 '***' 0.001 '**' 0.01 '*' 0.05 '.' 0.1 ' ' 1
##
## Approximate significance of smooth terms:
##                    edf Ref.df     F p-value
## s(Tmax26):HVIL 2.000  2.000 34.46  <2e-16 ***
## s(Tmax26):HVIM 2.879  3.311 79.35  <2e-16 ***
## s(Tmax26):HVIH 2.436  2.742 77.38  <2e-16 ***
## ---
## Signif. codes:  0 '***' 0.001 '**' 0.01 '*' 0.05 '.' 0.1 ' ' 1
##
## R-sq.(adj) =  0.897   Deviance explained = 90.7%
## GCV = 1.7225  Scale est. = 1.5467    n = 84

summary(quasiPoisHVIfitM3) ## GLM

##
## Family: quasipoisson
## Link function: log
##
## Formula:
## H ~ offset(log(PD)) + Tmax26 + I(Tmax26^2) + HVI + HVI:Tmax26
##
## Parametric coefficients:
##               Estimate Std. Error t value Pr(>|t|)
## (Intercept) -16.173263   0.286151 -56.520  < 2e-16 ***
## Tmax26        0.021512   0.044712   0.481 0.631801
## I(Tmax26^2)   0.008785   0.001772   4.958 4.13e-06 ***
## HVI1          0.174407   0.240086   0.726 0.469774
## HVI2          0.027227   0.214595   0.127 0.899368
## Tmax26:HVI1  -0.059510   0.016456  -3.616 0.000531 ***
## Tmax26:HVI2   0.002206   0.014413   0.153 0.878739
## ---
## Signif. codes:  0 '***' 0.001 '**' 0.01 '*' 0.05 '.' 0.1 ' ' 1
##
##
## R-sq.(adj) =  0.894   Deviance explained = 90.2%
## GCV = 1.6589  Scale est. = 1.5876    n = 84

## Practically no difference in deviance:
anova(quasiPoisHVIfitM3,quasiPoisgamHVIfit)
```

```
## Analysis of Deviance Table
##
## Model 1: H ~ offset(log(PD)) + Tmax26 + I(Tmax26^2) + HVI + HVI:Tmax
26
## Model 2: H ~ offset(log(PD)) + HVI + s(Tmax26, m = 3, by = HVI)
##   Resid. Df Resid. Dev     Df Deviance
## 1    77.000     117.09
## 2    72.947     111.33 4.0533   5.7586
```

##### Risk Derivatives by HVI Category

We evaluate a finite difference approximation to the derivative of $\mu(t, \beta)$ with respect to *t*, which is, by the chain rule,

$$\frac{\mathrm{d}\mu}{\mathrm{d}t} = \frac{\mathrm{d}\mu}{\mathrm{d}\eta}\frac{\mathrm{d}\eta}{\mathrm{d}t},$$

where the linear predictor $\boldsymbol{\eta} = \boldsymbol{h^t\beta}$, $\boldsymbol{h} = (h_1(t), \ldots, h_J(t))^t$ is the smooth (spline) function's J basis functions (transformations) evaluated at temperature *t,* and $\boldsymbol{\beta}$ are the corresponding parameters (not the same as our quasi-Poisson models' parameters, of course). When evaluated at the n observed temperatures, these basis functions give the typical $(n \times J)$`regression' matrix of the linear predictor. The code that we use for computing derivatives is a modified version of the code given in the example entitled "Differentiating the smooths in a model (with CIs for derivatives)," in the help entry for the predict.gam function in the mgcv R library package (34); see also Wood (35 pp. 341-343) on variances of non-linear functions of the linear predictor.

For each HVI group, this first chunk of code, below, computes the finite difference approximation to the derivative of the risk curve, $\mu(t, \beta)$, with respect to temperature, *t*, using the chain rule, as discussed above in this section.

```
## Mesh of temperature values on which to evaluate derivatives:
x.mesh <- seq(min(heat.df$Tmax26),
              max(heat.df$Tmax26),length=200)
## Use median (i.e., `typical') person-day offset for prediction (to
## be divided out in the end):
newd <- data.frame(PD=rep(median(heat.df$PD),200), Tmax26=x.mesh)
HVIL<- factor(rep("L", 200), levels=levels(heat.df$HVI))
HVIM<- factor(rep("M", 200), levels=levels(heat.df$HVI))
HVIH<- factor(rep("H", 200), levels=levels(heat.df$HVI))
newdL <- cbind.data.frame(newd, HVI=HVIL)
newdM <- cbind.data.frame(newd, HVI=HVIM)
newdH <- cbind.data.frame(newd, HVI=HVIH)
## Evaluate basis function matrix (i.e. `regression' matrix) on mesh
## (for typical person-day offset and by group):
X0L <- predict(quasiPoisgamHVIfit, newdL, type="lpmatrix")
X0M <- predict(quasiPoisgamHVIfit, newdM, type="lpmatrix")
X0H <- predict(quasiPoisgamHVIfit, newdH, type="lpmatrix")
## Finite difference interval:
eps <- 1e-7
```

```
## Shift the evaluation mesh:
x.mesh <- x.mesh + eps
## Evaluate basis function matrix (i.e. `regression' matrix) on
## shifted mesh:
newd<- data.frame(PD=rep(median(heat.df$PD),200), Tmax26=x.mesh + eps)
newdL <- cbind.data.frame(newd, HVI=HVIL)
newdM <- cbind.data.frame(newd, HVI=HVIM)
newdH <- cbind.data.frame(newd, HVI=HVIH)
X1L <- predict(quasiPoisgamHVIfit, newdL, type="lpmatrix")
X1M <- predict(quasiPoisgamHVIfit, newdM, type="lpmatrix")
X1H <- predict(quasiPoisgamHVIfit, newdH, type="lpmatrix")

## Xp is finite diff. approx. deriv. of basis function matrix wrt
## temperature for mesh:
XpL <- (X1L-X0L)/eps
XpM <- (X1M-X0M)/eps
XpH <- (X1H-X0H)/eps

## Xp%*%beta is finite diff. approx. deriv. of linear predictor,
## deta/dt, evaluated on mesh.  This is a component of the chain rule
## applied to the derivative of the risk function itself, obtained in
## subsequent code, dm/dt = dm/deta * deta/dt.
## detadtL<- XpL%*%coef(quasiPoisgamHVIfit)
## detadtM<- XpM%*%coef(quasiPoisgamHVIfit)
## detadtH<- XpH%*%coef(quasiPoisgamHVIfit) ## <-- see next chunk
```

##### Small Ensemble Illustration

Probability envelopes for the derivative curves are based on ensembles of curves.  A few ensemble member curves are plotted here for the HVI=high category as an illustration.  See Wood (2017, 341-343).

```
## Small ensemble of risk curves for HVI=high: m(t): dm/dt = dm/deta
## * deta/dt = exp(eta) * deta/dt. Note that eta omits (i.e., is
## adjusted for) offset but includes intercept (baseline).
set.seed(8675309 + 99)
betar<- rmvn(n=1000, beta<- coef(quasiPoisgamHVIfit),
             vcov(quasiPoisgamHVIfit))
## dm/dt = dm/deta * deta/dt = exp(eta) * deta/dt:
expetaL<- exp(X0L%*%t(betar)) ## exp(eta) low
expetaM<- exp(X0M%*%t(betar)) ## exp(eta) med
expetaH<- exp(X0H%*%t(betar)) ## exp(eta) high

detadtL<- XpL%*%t(betar) ## deta/dt low
detadtM<- XpM%*%t(betar) ## deta/dt med
detadtH<- XpH%*%t(betar) ## deta/dt high

dmdtL<- expetaL * detadtL ## dm/dt low
dmdtM<- expetaM * detadtM ## dm/dt med
dmdtH<- expetaH * detadtH ## dm/dt high
```

```
plot(x=range(heat.df$Tmax26)+26, y=range(dmdtH), type="n",
     ylab=expression("Risk Derivative"~hat(mu)*minute(t)),
     xlab="Temperature t")
apply(dmdtH, 2, function(x, mesh){
   lines(x=mesh+26, y =x, lty=3, col="grey")
}, mesh=x.mesh)

## add fitted deriv for HVI = high:
expetaH<- exp(X0H%*%beta) ## dm/deta = exp(eta) high
detadtH<- XpH%*%beta ## deta/dt high
dmdtH<- expetaH * detadtH ## dm/dt = dm/deta * deta/dt high
lines(x=x.mesh+26, y=dmdtH)

abline(h=0, col="black")
title(main="Risk Derivative Ensemble", sub="HVI=high")
```

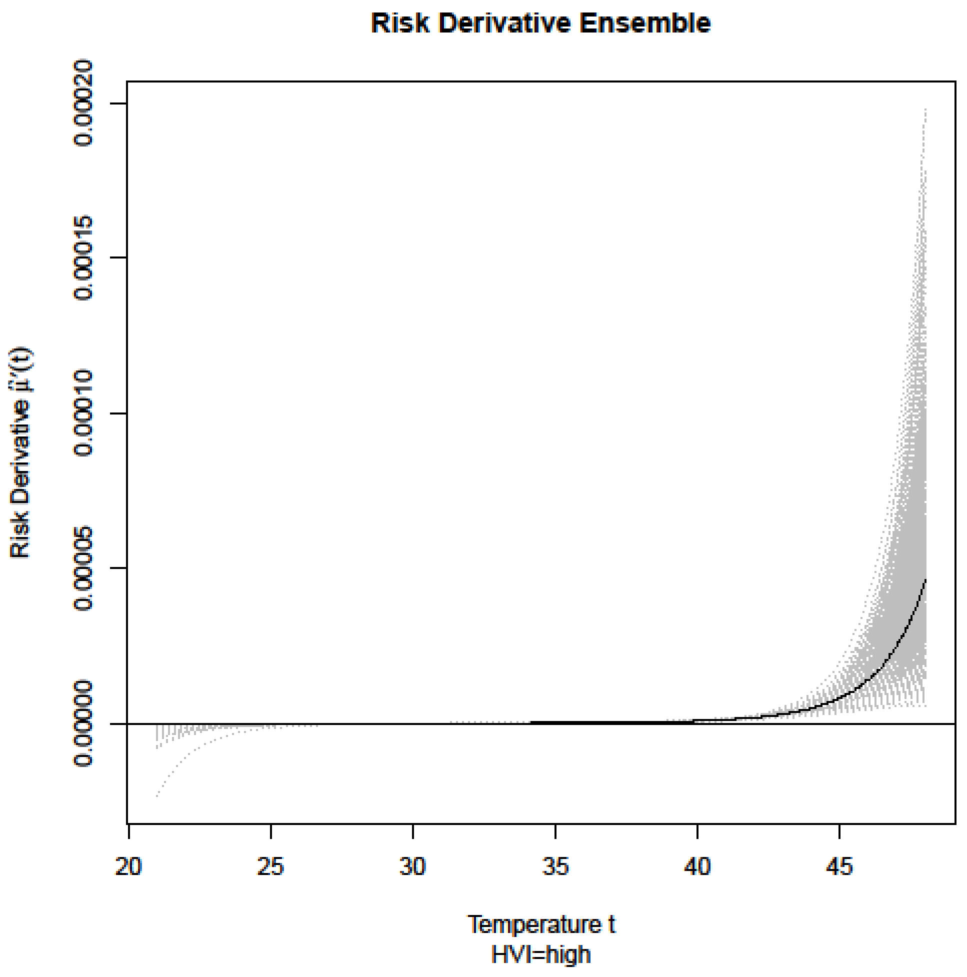


**Supplemental Figure S2. Risk Derivative Ensemble for HVI=High Vulnerability Category**

**Point-wise 98% Probability Envelopes of Risk Derivative**

The following figure, Supplemental Figure 3, uses a much larger ensemble than the one shown above to compute a 98% point-wise probability envelopes for the risk derivative curves for each HVI category. We see all three risk derivatives statistically (and visually) coincide at $t_{max}$=26 degrees and zero derivative value (vertical and horizontal solid lines), and we choose tmax=26 as our baseline temperature in the quasi-Poisson GLM (Model 3) used throughout the main text.

```
set.seed(8675309 + 99)
betar<- rmvn(n=100000, coef(quasiPoisgamHVIfit), vcov(quasiPoisgamHVIfi
t))
## dm/dt = dm/deta * deta/dt = exp(eta) * deta/dt
expetaL<- exp(X0L%*%t(betar))
expetaM<- exp(X0M%*%t(betar))
expetaH<- exp(X0H%*%t(betar))

detadtL<- XpL%*%t(betar)
detadtM<- XpM%*%t(betar)
detadtH<- XpH%*%t(betar)

dmdtL<- expetaL * detadtL
dmdtM<- expetaM * detadtM
dmdtH<- expetaH * detadtH

meancurveL<- rowMeans(dmdtL)
meancurveM<- rowMeans(dmdtM)
meancurveH<- rowMeans(dmdtH)

bL<- apply(dmdtL, 1, quantile, probs=c(0.01,0.99))
bM<- apply(dmdtM, 1, quantile, probs=c(0.01,0.99))
bH<- apply(dmdtH, 1, quantile, probs=c(0.01,0.99))

zoomindx<- which((x.mesh >= 0) & (x.mesh <= 8))
plot(x=c(0,8)+26, y=range(cbind(bL[,zoomindx],bM[,zoomindx],bH[,zoomind
x])),
     type="n", ylab=expression("Risk Derivative"~hat(mu)*minute(t)~"and
bounds"),
     xlab="Temperature t")
lines(x=x.mesh+26, y=meancurveL, lwd=1.5, lty=2)
lines(x=x.mesh+26, y=meancurveM, lwd=1.5, lty=3)
lines(x=x.mesh+26, y=meancurveH, lwd=1.5, lty=4)

lines(x=x.mesh+26, y=bL[1,], lty=2, col="grey")
lines(x=x.mesh+26, y=bM[1,], lty=3, col="grey")
lines(x=x.mesh+26, y=bH[1,], lty=4, col="grey")

lines(x=x.mesh+26, y=bL[2,], lty=2, col="grey")
lines(x=x.mesh+26, y=bM[2,], lty=3, col="grey")
lines(x=x.mesh+26, y=bH[2,], lty=4, col="grey")
```

```
abline(h=0, v=30, col="black", lty=1)
legend("bottomright", legend=c("high","med","low","zero"),
       lty=c(4,3,2,1),
       title="HVI Category", cex=0.8)
title(main="98% Point-wise Probability Bounds \n for Risk Derivatives")
```

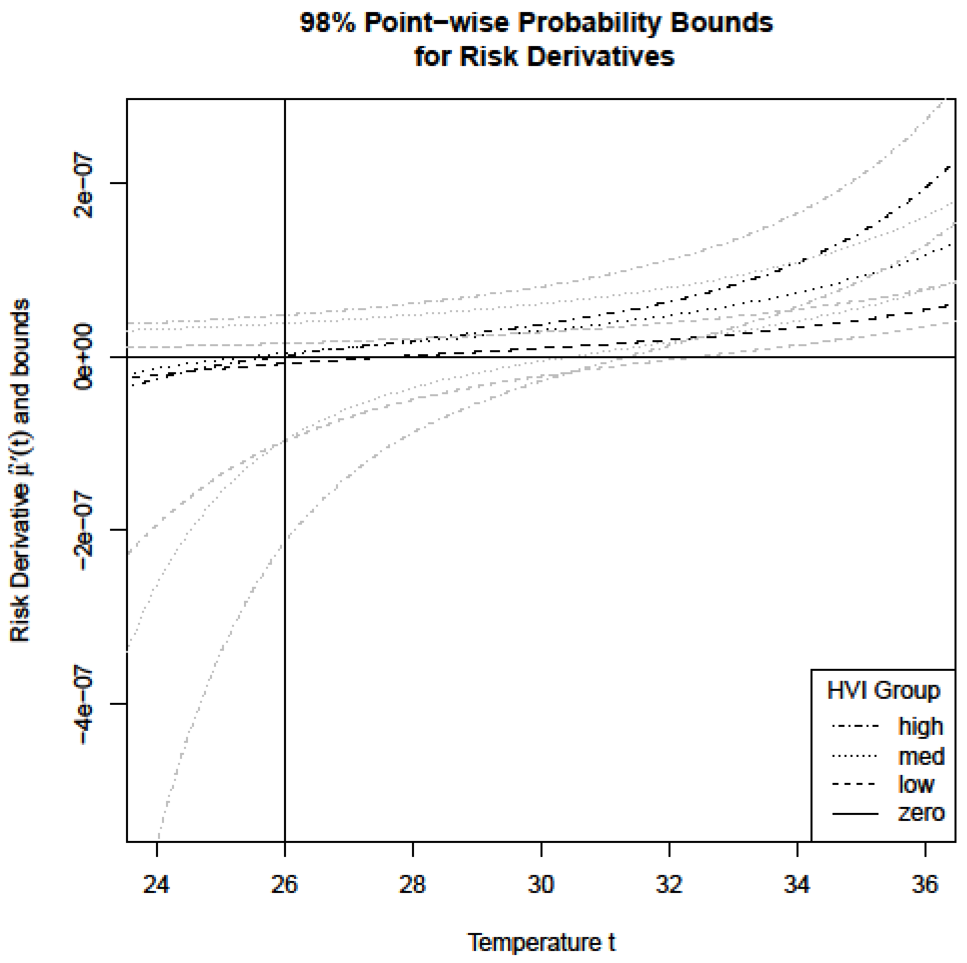


**Supplemental Figure S3.  98% Point-wise Probability Bounds for Risk Derivatives**

A Socio-spatial Model of the Risk of Hospitalization from Vulnerability to High Temperatures

#### Supplemental Information. Section 3. Detail on Person-days of Exposure and Number of Heat-related Hospitalizations

Supplemental Table S2 provides detail on the number of person-days of exposure and the number of heat-related hospitalization for increasing maximum temperature by category of Heat Vulnerability Index. Note that for each row, the sum of heat-related hospitalizations in Low, Medium, and High HVI categories does not equal the overall number of hospitalizations in the same row. The total number of hospitalizations for *all* rows does. The reason is that for the HVI categories, $t_{max}$ was assigned using the closest weather station (as described in the Temperature subsection of the Methods section in the main text), whereas for the Overall, $t_{max}$ was assigned using the Phoenix Sky Harbor station. Note that hospitalizations were observed at $t_{max=}$ 47 and $t_{max}$=48, but we did not report these these in Figure 1 in the main text due to the small number of cases.

**Supplemental Table S2. Number of Person-Days of Exposure and Number of Heat-Related Hospitalization for Increasing Maximum Temperature ($t_{max}$) by Category of Heat Vulnerability Index. Maricopa County 2005-2009 Heat-Related Hospitalization Analysis.**

| | Category of Heat Vulnerability Index | | | | | | | |
|---|---|---|---|---|---|---|---|---|
| | Low | | Medium | | High | | Overall | |
| Tmax (°C) | Number of Person-Days | Number of Heat-Related Hospitalizations | Number of Person-Days | Number of Heat-Related Hospitalizations | Number of Person-Days | Number of Heat-Related Hospitalizations | Number of Person-Days | Number of Heat-Related Hospitalizations |
| 21 | 527,679 | 0 | 1,010,168 | 1 | 350,882 | 0 | 1,888,729 | 1 |
| 22 | 1,810,305 | 0 | 1,614,083 | 0 | 400,990 | 0 | 3,825,378 | 0 |
| 23 | 1,813,709 | 0 | 1,773,183 | 0 | 436,947 | 0 | 4,023,839 | 0 |
| 24 | 3,066,962 | 0 | 3,785,704 | 0 | 1,475,028 | 0 | 8,327,694 | 0 |
| 25 | 5,558,489 | 1 | 7,154,126 | 0 | 2,619,208 | 0 | 15,331,823 | 1 |
| 26 | 9,957,177 | 2 | 10,525,379 | 0 | 3,596,076 | 0 | 24,078,632 | 2 |
| 27 | 10,394,010 | 1 | 11,049,916 | 2 | 3,719,670 | 0 | 25,163,596 | 3 |
| 28 | 13,429,149 | 0 | 16,977,117 | 1 | 6,552,022 | 1 | 36,958,288 | 2 |
| 29 | 12,541,766 | 2 | 12,870,329 | 0 | 4,442,190 | 1 | 29,854,285 | 3 |
| 30 | 21,394,730 | 1 | 23,156,115 | 2 | 8,642,720 | 0 | 53,193,565 | 3 |
| 31 | 25,451,815 | 2 | 27,523,749 | 4 | 10,063,231 | 2 | 63,038,795 | 8 |
| 32 | 30,448,029 | 5 | 30,769,234 | 7 | 10,623,925 | 5 | 71,841,188 | 17 |
| 33 | 35,890,154 | 5 | 39,573,084 | 8 | 15,080,690 | 1 | 90,543,928 | 14 |
| 34 | 28,477,480 | 7 | 34,405,204 | 9 | 13,079,309 | 7 | 75,961,993 | 23 |
| 35 | 45,841,121 | 5 | 50,278,495 | 6 | 18,465,043 | 3 | 114,584,659 | 14 |
| 36 | 59,044,855 | 11 | 60,148,287 | 16 | 22,329,658 | 9 | 141,522,800 | 36 |
| 37 | 56,681,680 | 14 | 63,769,989 | 38 | 24,321,088 | 13 | 144,772,757 | 65 |
| 38 | 59,467,911 | 16 | 64,172,166 | 36 | 23,714,136 | 19 | 147,354,213 | 71 |
| 39 | 49,344,786 | 30 | 56,949,531 | 36 | 21,638,296 | 27 | 127,932,613 | 93 |
| 40 | 67,819,292 | 10 | 71,257,294 | 44 | 27,597,067 | 32 | 166,673,653 | 86 |
| 41 | 53,006,881 | 23 | 59,384,513 | 56 | 21,800,408 | 29 | 134,191,802 | 108 |
| 42 | 52,579,458 | 24 | 51,627,623 | 55 | 17,889,599 | 51 | 122,096,680 | 130 |
| 43 | 33,442,763 | 26 | 34,848,904 | 62 | 11,914,543 | 34 | 80,206,210 | 122 |
| 44 | 16,333,981 | 26 | 16,135,275 | 56 | 4,894,432 | 36 | 37,363,688 | 118 |
| 45 | 9,718,595 | 8 | 7,612,247 | 20 | 2,035,644 | 12 | 19,366,486 | 40 |
| 46 | 1,771,975 | 4 | 2,345,607 | 17 | 749,323 | 14 | 4,866,905 | 35 |
| 47 | 660,658 | 1 | 480,564 | 4 | 110,134 | 4 | 1,251,356 | 9 |
| 48 | 42,151 | 0 | 94,879 | 2 | 32,537 | 1 | 169,567 | 3 |
| Total | 706,517,561 | 224 | 761,292,765 | 482 | 278,574,796 | 301 | 1,746,385,122 | 1,007 |

A Socio-spatial Model of the Risk of Hospitalization from Vulnerability to High Temperatures

#### Supplemental Information. References


1. Reid CE, O'Neill MS, Gronlund CJ, Brines SJ, Brown DG, Diez-Roux AV, et al. Mapping community determinants of heat vulnerability. Environmental Health Perspectives. 2009;117(11):1730–1736.
2. Reid CE, Mann JK, Alfasso R, English PB, King GC, Lincoln RA, et al. Evaluation of a Heat Vulnerability Index on abnormally hot days: An environmental public health tracking study. Environmental Health Perspectives. 2012;120(5):715–720.
3. Harlan SL, Declet-Barreto JH, Stefanov WL, Petitti DB. Neighborhood effects on heat deaths: social and environmental predictors of vulnerability in Maricopa County, Arizona. Environmental Health Perspectives. 2013;121(2):197-204.
4. Klinenberg E. *Heat Wave: A Social Autopsy of Disaster in Chicago*. University of Chicago Press; Chicago. 2002.
5. O'Neill MS, Zanobetti A, Schwartz J. Disparities by race in heat-related mortality in four US cities: the role of air conditioning prevalence. Journal of Urban Health. 2005;82(2):191–197.
6. Chakalian PM, Kurtz L, Harlan SL, White D, Gronlund CJ, Hondula DM. Exploring the social, psychological, and behavioral mechanisms of heat vulnerability in the city of Phoenix, AZ. Journal of Extreme Events. 2019;6(03n04):2050006.
7. Watkins LE, Wright MK, Kurtz LC, Chakalian PM, Mallen ES, Harlan SL, et al. Extreme heat vulnerability in Phoenix, Arizona: a comparison of all-hazard and hazard-specific indices with household experiences. Applied Geography. 2021;102430.
8. McMichael AJ, Woodruff RE, Hales S. Climate change and human health: present and future risks. The Lancet. 2006;367(9513):859–869.
9. Yip F, Flanders W, Wolkin A, Engelthaler D, Humble W, Neri A, et al. The impact of excess heat events in Maricopa County, Arizona: 2000-2005. International Journal of Biometeorology. 2008;52(8):765–772.
10. Bouchama A, Dehbi M, Mohamed G, Matthies F, Shoukri M, Menne B. Prognostic factors in heat wave–related deaths: a meta-analysis. Archives of Internal Medicine. 2007;167(20):2170–2176.
11. Lochner KA, Kawachi I, Brennan RT, Buka SL. Social capital and neighborhood mortality rates in Chicago. Social Science & Medicine. 2003;56(8):1797–1805.

28. United States Census Bureau. 2010 Census of Population and Housing. 2010.

29. United States Census Bureau. 2006-2010 American Community Survey 5-year Estimates. United States Census Bureau; 2010.

30. Maricopa County Assessor’s Office. GIS Parcels Database. 2010.

31. Tucker CJ. Red and photographic infrared linear combinations for monitoring vegetation. Remote Sensing of Environment. 1979;8(2):127–150.

32. NASA Landsat Program. Landsat TM scene L5037037_03720100813 [Internet]. USGS, Sioux Falls, 8/13/2010; 2010. Available from: http://landsat.gsfc.nasa.gov/

33. Wood SN. Thin plate regression splines. Journal of the Royal Statistical Society: Series B (Statistical Methodology). 2003;65(1):95–114.

34. Wood SN. Fast stable restricted maximum likelihood and marginal likelihood estimation of semiparametric generalized linear models. Journal of the Royal Statistical Society: Series B (Statistical Methodology). 2011;73(1):3–36.

35. Wood SN. *Generalized Additive Models: An Introduction with R*. *2$^{nd}$ Edition.* CRC Press; Boca Raton. 2017.
